# Socioeconomic disparities in perceived air quality and associated respiratory health outcomes among residents of Nairobi, Kenya

**DOI:** 10.64898/2026.08.19.26360866

**Authors:** Eunice Otieno, James Mwitari, George Makalliwa

## Abstract

The socioeconomic inequality in exposure to air pollution particularly, the fine particulate matter with an aerodynamic diameter of 2.5 micrometer (PM_2.5_) possess a significant public health challenge, yet little is known about how the disparities vary across the various economic status areas in Nairobi. Globally, studies have shown that exposure to air pollution is unequal across communities, leading to disparities in harm to human health.

This study examined the association between socioeconomic characteristics and perceived air quality among residents of low- and high-socioeconomic status areas in Nairobi, Kenya.

Two regions within Nairobi County were selected for this study: Mukuru kwa Njenga (representing the Low Socioeconomic Status) and Lang’ata (representing the High Socioeconomic Status) with a sample size of 384 in HSES areas and 368 in LSES areas.

A cross-sectional study was conducted among 752 respondents residing in selected LSES and HSES areas of Nairobi. Data was collected using a structured questionnaire assessing sociodemographic characteristics, income, education, employment, perceived air quality, and self-reported health outcomes associated with air pollution exposure. Descriptive statistics were used to summarize participant characteristics and perceived air quality. Chi-square tests were used to examine associations between residential area and categorical health outcomes, while ordinal logistic regression was used to assess the association between socioeconomic characteristics and perceived air-quality ratings.

Perceived air quality differed significantly between residential socioeconomic groups. Respondents in LSES areas were substantially more likely to rate air quality as poor or very poor, with 45.4% rating it as very poor, compared with only 1.6% of respondents in HSES areas. In contrast, 12.2% of HSES respondents rated air quality as good compared with 0.3% in LSES areas. The association between area of residence and perceived air-quality rating was statistically significant, χ²(3) = 282.672, p < 0.001. In the ordinal logistic regression model, HSES residence was associated with significantly lower odds of reporting poorer perceived air quality compared with LSES residence (OR = 0.135, 95% CI: 0.095–0.190, p < 0.001). Income was also significantly associated with perceived air quality, while respondents with no formal education had higher odds of reporting poorer perceived air quality compared with those with secondary education (OR = 3.254, 95% CI: 1.388–7.638, p = 0.007).

Significant differences were also observed for several self-reported health outcomes. Respiratory problems were more prevalent among respondents in LSES areas than HSES areas (72.7% versus 50.4%; χ²(1) = 29.081, p < 0.001; Cramer’s V = 0.224). However, allergies, eye irritation, and headaches were reported more frequently in HSES areas than in LSES areas, with significant associations observed for allergies (χ²(1) = 106.479, p < 0.001; Cramer’s V = 0.429), eye irritation (χ²(1) = 136.577, p < 0.001; Cramer’s V = 0.486), and headaches (χ²(1) = 149.180, p < 0.001; Cramer’s V = 0.508). No statistically significant association was observed for cardiovascular problems, likely reflecting the very low number of reported cases.

Substantial socioeconomic disparities in perceived air quality and self-reported respiratory health outcomes were observed. Residents of low-socioeconomic status areas consistently perceived poorer air quality and reported a higher burden of respiratory problems, highlighting the need for targeted interventions to reduce environmental health inequalities.

## INTRODUCTION

### Background Information

Air pollution has the greatest impact on human health of any environmental health risk. It harms health and kills in much the same way as smoking: by increasing the risk of developing cardiovascular and respiratory diseases, and lung cancer (World Health Organization, 2014). Exposure to polluted air is now recognized as a vital risk factor for noncommunicable human disease conditions (Schraufnagel et al., 2019). Moreover, a quarter of the world population is exposed to high concentrations of air pollution (Landrigan, 2017).

Air pollution constitutes a wide range of pollutants. However, of particular concern is the fine particulate matter with an aerodynamic diameter of 2.5 micrometer (PM_2.5_). PM_2.5_ has been associated with several cardiovascular and respiratory complications (Hicken et al., 2023). PM_2.5_ concentration in the atmosphere is characterized by several activities such as electronic waste recycling, heavy car traffic, and industrial processes (Geng et al., 2021). Majority of these activities associated with emission of air pollutants are within urban settings.

A study by Cohen et al. (2017) reported that about 4.2 million deaths globally were as a result of exposure to PM_2.5_. Ambient air pollution was among the leading causes of mortality in 2016 in the United (US) (Murray et al., 2018). Additionally, exposure to air pollution was reported to be higher among ethnic minorities and those of low socioeconomic status in the US (Huang et al.,2019; Lamichhane et al., 2020; Bravo et al., 2022). Globally, studies have shown that exposure to air pollution is unequal across communities, leading to disparities in harm to human health.

WHO (2014) reported that health risks associated with air pollution are higher in low- and middle-income countries, where 88% of the premature deaths are due to air pollution. In a related study, Ngo et al. (2018) reported that air pollution levels are usually higher in the most deprived areas within a city. If unchecked, the impact of air quality on health will widen the inequality gap between the low-income earners and the high-income earners.

In Africa, recent evidence increasingly demonstrates that air pollution exposure is socially differentiated, with disadvantaged populations often experiencing a disproportionate burden of exposure and associated health risks. In South Africa, a recent systematic review found that socioeconomic disadvantage, poverty, informal and low-quality housing, proximity to industrial activities, and dependence on polluting household fuels contribute to heightened exposure among vulnerable communities (Mdluli et al., 2025). In Kenya, emerging evidence indicates that air-quality disparities are closely associated with socioeconomic conditions, urban spatial inequality, and unequal access to cleaner household energy (Egondi et al. 2018). Additionally, households in informal settlements and other low-income communities experience particularly high levels of air pollution because of unpaved roads and open burning of waste.

Nairobi is one of the fastest growing cities in Africa. The growth has been characterized by increase in industries and vehicles (deSouza et al., 2021). However, the growth has seen increase in air pollution due to poor waste management practices such as open burning, vehicular and industrial emissions. Noteworthy, most of Nairobi’s population live in the slums with poor social services, hence more exposed to air pollution. According to a 2017 UNEP study, Nairobi’s air breaches all limits set by the WHO. West et al. (2020) and Egondi et al. (2018) reported that air pollution is a major concern in Nairobi’s poor neighborhoods. Concerns on air quality in Nairobi led to the development of Nairobi City County Air Policy, 2020 and an Air Quality Action Plan. Recognizing the burden of air pollution on public health, the National Environment Management Authority (NEMA) imposed regulations in 2014 (revised in 2024) for national air quality standards. The regulations laid out steps to be undertaken for the prevention, control and abatement of air pollution. These initiatives demonstrate efforts by the National and County governments to improve air quality and protect public health.

Despite the existence the regulatory and policy frameworks, there has been little or no enforcement of air quality regulations in most areas. In addition, most studies on air quality disparities and their association with health outcomes have been conducted in developed countries particularly the United States of America (Penza et al., 2014, Sun et al.,2016: Mueller et ak.,2020). Empirical studies assessing air quality disparities between LSES areas and HSES areas in Nairobi are Limited. Consequently, it is not clear whether the burden of air pollution is homogenous or varies across the different socioeconomic status areas within Nairobi.

This study examined the association between socioeconomic characteristics and air quality among residents of low- and high-socioeconomic status areas in Nairobi. Information obtained from this study can be used by stakeholders and government agencies to develop and implement effective policies and interventions for the management of air pollution, promote environmental justice and ensure improvement in air quality and health outcomes among Nairobi residents.

## MATERIALS AND METHODS

### Study Area

The study was conducted in Nairobi, Kenya’s capital city, and one of the fastest growing cities in Africa. The county’s population in 2022 was 5,598,338 (Kenya National Bureau of Statistics (KNBS), 2020), with about 58% of the population living in the informal settlements (Beguy et al., 2015). Two regions were selected for this study: Mukuru kwa Njenga (representing the LSES) and Lang’ata (representing the HSES).

Mukuru Kwa Njenga, Mukuru Kwa Reuben, and Viwandani (together referred to as Mukuru) are three adjoining informal communities in Nairobi’s Embakasi South and Makadara subcounties. They are situated on a mix of privately and publicly held land. This area is home to over 100,000 households, most of whom live in small, overcrowded shacks made of sheet metal with concrete or earth floors. The availability of services is quite limited. Sanitation is inadequate, waste disposal is poorly handled, open drainage channels are common, and access to electricity and water sources is limited. The community’s multiple burdens present themselves in severe health concerns, including high levels of respiratory disease (Gulis et al., 2004) and cardiovascular disease, which are major causes of death locally (Mberu et al., 2015).

Langata sub County is one of the 17 administrative units of Nairobi City County (NCC) with an estimated population of 176,314 people living in 52,656 households occupying an area of 196.80 km2. It is bounded to the North by Dagoretti sub County, Kibra sub County, Starehe sub County, Embakasi sub County, Machakos County and to the South by Kajiado County. It is about 10 km to the South of Central Business District. It has five wards, namely; Karen, Mugomoini, Nairob-West, South C and Nyayo High-Rise.

### Study Design

Cross-sectional design was used to assess the disparities in socioeconomic status and health outcomes. Recruitment of participants and data collection was carried out from 16/09/2025 to 20/11/2025. Self-administered questionnaires were used to understand people’s socioeconomic status, perceptions toward air pollution and the perceived health impacts.

### Study Population

The study involved residents of Mukuru and Lang’ata Sub-Counties within Nairobi County. The target population were those who had lived in the selected areas for more than three years.

### Inclusion Criteria

To be included into the study, the participants were required to be above 18 years of age, be able to consent to study participation, and having lived in the study area for more than three years.

### Exclusion Criteria

Individuals who did not meet the inclusion criteria were excluded from this study. This included individuals below 18 years, individuals who had lived in the area for less than three years, and those who did not consent to participate in the study.

### Study Variables

The study variables were categorized into independent and dependent variables.The independent variable was the socioeconomic status (SES) of the study areas which were categoried as LSES areas and HSES areas. This variable included demographic and household characteristics of the respondents such as age, sex, level of education, occupation and income level. The dependent variables were the perceived health outcomes.

### Data Collection Tools

Primary data was collected using closed and open ended questionnaires, which were self-administered. The primary data included the demographic characteristics of the respondents, perception on air quality, perceived sources of air pollution and perceived health outcomes.

### Pretesting of Data Collection Tools

The questionnaire was pre-tested among a small, representative sample of the target population to assess clarity, relevance, comprehensibility, and cultural appropriateness. Feedback was used to identify and address ambiguities, inappropriate response options, language issues, and potential cultural sensitivities. Where applicable, the questionnaire was translated into the relevant local language and back-translated to ensure consistency. The reliability of multi-item measures was assessed using Cronbach’s alpha, while the pre-testing process helped establish the suitability and validity of the questionnaire for the study population.

### Data Collection

Data on socioeconomic and perception on health outcome was collected by use of self-administered questionnaires through the help of trained enumerators.

### Data Management and Analysis

Raw data collected from the field using questionnaires was cleaned to remove any incomplete data in Microsoft Excel. Thereafter, the data was coded based on the study parameters, and categorized as per the different socioeconomic classes. Quantitatve data collected using the questionnaire was analysed using the Statistical Package for Social Sciences (SPSS V.25). Perceived air quality was measured using an ordinal four-category variable: very poor, poor, fair, and good. The categories were coded from 1 to 4, respectively, with higher scores representing more favourable perceived air quality. Because the dependent variable was ordinal, an ordinal logistic regression model with a cumulative logit link was fitted to assess the association between socioeconomic characteristics and perceived air quality.

### Ethical Considerations

Prior to commencement of the study, ethical approval was obtained from the Institutional Scientific Ethics Review Committee, University of Eastern Africa, Baraton, approval number UEAB/ISERC/11/09/2024 and the National Commission for Science, Technology and Innovation (NACOSTI) license number NACOSTI/P/24/414593. Additional approval was obtained from the Nairobi County Commissioners office, approval reference no. ED 10/6VOL.XXIX (67). The study included adults aged 18 years and above, and no minors were recruited. Participation was voluntary, and written informed consent was obtained from all participants before enrolment in the study. Participants were informed about the purpose and procedures of the study, their right to decline to answer any question they felt uncomfortable with, and their right to withdraw from the interview at any time without penalty. Information obtained from participants was treated confidentially and was used solely for the purposes of this study.

## RESULTS

### Influence of Socioeconomic Factors on Air Quality Exposure

### Socio-demographic characteristics by Area of Residence

The planned sample size was 768 participants, with 384 participants allocated to each socioeconomic group. However, the final sample comprised 752 participants, including 384 from HSES areas and 368 from LSES areas. The 16-participant shortfall in the LSES group was due to incomplete questionnaires that were excluded from the final analysis. The final sample represented 97.9% of the planned sample size. Although this reduction may have resulted in a slight decrease in statistical power, the relatively small shortfall is unlikely to have substantially affected the study’s ability to detect meaningful associations, given the large overall sample size.

In terms of gender, both areas were female-dominated, however, this was more pronounced in the LSES area, where females constituted 84.2% of respondents compared to 61.2% in HSES. This may reflect higher participation of women in household-level surveys, particularly in informal settlements. Age distribution shows that LSES respondents were generally younger, with a higher proportion in the 18–30 age group (35.9%) compared to HSES (19.5%). Conversely, HSES areas had a higher proportion of older respondents, particularly in the 31–60 age brackets. This suggests differences in population structure, with LSES areas characterized by a more youthful demographic.

Educational attainment also differed notably between the two areas. While secondary education was the most common level in both settings, it was more prevalent in HSES (64.1%) than in LSES (46.7%). LSES areas had a higher proportion of respondents with only primary education or no formal education, indicating comparatively lower educational attainment. Employment patterns further highlight socioeconomic disparities. Most respondents in HSES areas were self-employed (68.0%), whereas LSES areas had a significantly higher proportion of unemployed individuals (67.9%). This suggests more limited economic opportunities in LSES settings and a greater reliance on informal or unstable income sources.

Income distribution reinforces these differences. In HSES areas, most respondents fell within the low-income category (65.9%), with a notable proportion in the medium-income group (16.7%). In contrast, LSES areas were dominated by very low-income earners (66.3%), with very few respondents in higher income categories. This indicates a substantially lower economic status in LSES areas, consistent with their classification. Table 1 shows detailed socio-demographic characteristics of the respondents.

**Table 1:** Socio-demographic characteristics of the respondents by Area.

| Variable | Category | HSES n (%) | LSES n (%) |
| --- | --- | --- | --- |
| Respondents |  | 384 (51.1) | 368 (48.9) |
| **Gender** | Male | 149 (38.8) | 58 (15.8) |
|  | Female | 235 (61.2) | 310 (84.2) |
| **Age (years)** | 18–30 | 75 (19.5) | 132 (35.9) |
|  | 31–45 | 152 (39.6) | 126 (34.2) |
|  | 46–60 | 145 (37.8) | 97 (26.4) |
|  | >60 | 12 (3.1) | 13 (3.5) |
| **Education** | No formal | 10 (2.6) | 14 (3.8) |
|  | Primary | 95 (24.7) | 133 (36.1) |
|  | Secondary | 246 (64.1) | 172 (46.7) |
|  | College/University | 33 (8.6) | 49 (13.3) |
| **Employment** | Formal | 19 (4.9) | 22 (6.0) |
|  | Self-employed | 261 (68.0) | 96 (26.1) |
|  | Unemployed | 104 (27.1) | 250 (67.9) |
| **Income** | High (USD 10–19/day) | 4 (1.0) | 1 (0.3) |
|  | Medium (USD 5–9/day) | 64 (16.7) | 23 (6.3) |
|  | Low (USD 1–4/day) | 253 (65.9) | 100 (27.2) |
|  | Very low (< USD 1/day) | 63 (16.4) | 244 (66.3) |

### Sources of Air pollution

The most reported source of air pollution was open burning of waste (78.5%), followed by fuel used for cooking (64.8%) and dust from unpaved roads (53.3%) (Table 2). In contrast, traffic/vehicle emissions (19.2%) and industrial emissions (18.6%) were less frequently identified by respondents. The total percentage of 234.4% reflects the multiple-response nature of the question, whereby respondents could identify more than one source of air pollution; therefore, the percentages represent the proportion of respondents selecting each source.

**Table 2:** Reported sources of air pollution.

| Source of Air Pollution | Frequency (N) | Percent (%) | Multiple-response (%) |
| --- | --- | --- | --- |
| Traffic/Vehicle Emissions | 132 | 8.2 | 19.2 |
| Industrial Emissions | 128 | 8.0 | 18.6 |
| Dust from unpaved roads | 366 | 22.7 | 53.3 |
| Open burning of waste | 539 | 33.5 | 78.5 |
| Fuel used for cooking | 445 | 27.6 | 64.8 |
| **Total** | 1610 | 100.0 | *234.4 |

\*represents a multiple-response question whereby respondents could identify more than one source of air pollution, hence percentage is more than 100.

These results show that localized and household-level pollution sources dominate the study area. Open waste burning and cooking fuels are typically associated with limited waste management systems, reliance on biomass or low-quality fuels and informal settlement conditions. The prominence of dust from unpaved roads further indicates infrastructure-related pollution, especially in areas with poor road surfacing. Interestingly, traffic and industrial emissions were reported less frequently, suggesting that respondents may perceive immediate, visible, or nearby pollution sources more strongly than broader urban or industrial contributors. This highlights the importance of perception-based exposure, which may differ from measured pollution levels.

### Association Between Area of Residence and Socioeconomic factors

#### A. Income level

To further examine socioeconomic disparities between the study areas, a cross-tabulation analysis was conducted between area of residence and income level. The crosstabulation reveals a highly significant association between area of residence and income level (Pearson χ² = 193.897, df = 3, p < .001; Cramer’s V = .508, indicating a strong effect) as shown in Table 3. In HSES areas, 65.9% of respondents fell into the Low-income category and only 16.4% into Very low income. In contrast, LSES areas had a higher proportion of Very low-income respondents (66.3%) and far fewer in the Low-income bracket (27.2%). High- and Medium-income groups were mainly concentrated in HSES areas.

**Table 3:** Cross tabulation between area of residence and income level.

| Statistic | Value |
| --- | --- |
| Pearson Chi-Square | 193.897<sup>a</sup> |
| df | 3 |
| p- value | <.001 |
| Cramer's V | .508 |

This result demonstrates socioeconomic gradient underlying the area-based disparity: LSES areas are disproportionately inhabited by the poorest households. Lower-income residents in LSES areas are more likely to rely on polluting fuels such as charcoal, wood, kerosene, and live near waste dumps or unpaved roads, amplifying their exposure to air pollutants. The strong Cramer’s V confirms that income is a major driver of the observed HSES–LSES air quality divide. These findings align with global evidence showing that lower-SES communities consistently experience higher ambient and household air pollution exposure. In Nairobi slums, similar patterns have been documented where poverty concentrates households near pollution hotspots (West et al., 2020).

#### B. Education level

A statistically significant but moderate association exists between area of residence and education level (Pearson χ² = 22.892, df = 3, p < .001; Cramer’s V = .174) (Table 4). HSES areas had higher proportions of College/University (8.6%) and Secondary School (64.1%) respondents, while LSES areas showed higher Primary School (36.1%) and No Formal Education (3.8%) levels.

**Table 4:**
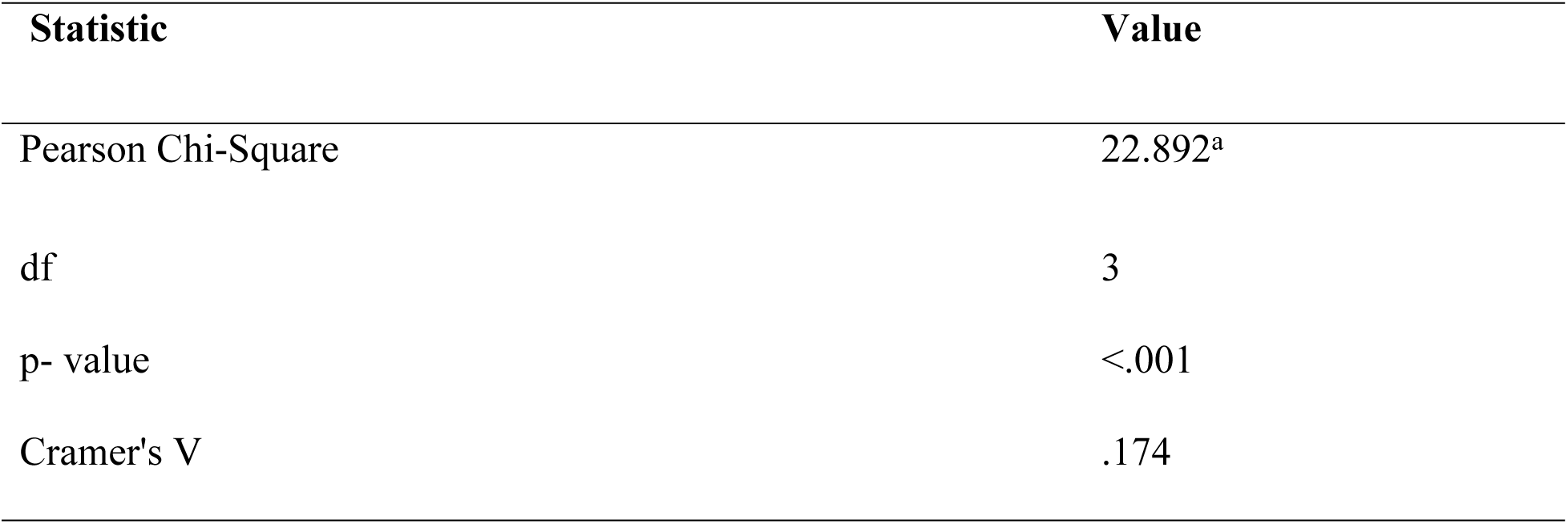
Association between area of residence and education level Statistic Value.

The results indicate that educational level reinforces the spatial disparity in air quality exposure. Lower education in LSES areas may limit awareness of pollution risks and adoption of cleaner practices, perpetuating higher exposure. The moderate effect size suggests education operates alongside income but is a secondary driver compared with the area itself. Dianati et al. (2019) in their study reported that lower educational attainment in informal settlements correlates with poorer environmental health perceptions and higher reliance on polluting fuels.

### Logistic Regression Analysis of Factors Associated with Air Quality

A logistic regression analysis was conducted to examine the influence of socioeconomic factors and area of residence on perceived air quality. The model included area of residence, income level, and education level as predictors. The ordinal logistic regression model (link = logit) was significant overall (χ² = 368.617, df = 7, p < .001; Nagelkerke R² = .423) as shown in Table 5.

**Table 5:** Ordinal Logistic Regression (Education + Income + Area)

| Model Fitting Information |  |  |  |  |  |
| --- | --- | --- | --- | --- | --- |
| Model | -2 Log Likelihood | Chi-Square | df | Sig. | Nagelkerke $R^2$ |
| Final | 319.773 | 368.617 | 7 | <.001 | .423 |
| Link function: Logit. |  |  |  |  |  |

The results indicate that area of residence was a significant predictor of air quality, with respondents in HSES areas significantly less likely to report poorer air quality compared to those in LSES areas (β = -2.005, p < 0.001) (Table 6). Income level was also significantly associated with air quality. Respondents in the low-income (β = -1.330, p < 0.001) and medium-income categories (β = -2.135, p < 0.001) were significantly less likely to report very poor air quality compared to those in the very low-income category. In terms of education, only respondents with no formal education showed a significant association (β = 1.180, p = 0.007), indicating a higher likelihood of reporting poorer air quality compared to those with secondary education. Other education categories were not statistically significant.

**Table 6:** Ordinal Logistic Regression Predicting Air-Quality Rating.

| Predictor | $\beta$ | SE | OR | 95% CI for OR | p-value |
| --- | --- | --- | --- | --- | --- |
| **Education** |  |  |  |  |  |
| College/University vs Secondary | -0.413 | 0.250 | 0.662 | 0.405–1.080 | .099 |
| No formal education vs Secondary | 1.180 | 0.435 | 3.254 | 1.388–7.638 | .007 |
| Primary vs Secondary | -0.234 | 0.167 | 0.791 | 0.571–1.096 | .159 |
| **Income** |  |  |  |  |  |
| High vs Very low | -1.236 | 0.911 | 0.290 | 0.049–1.733 | .175 |
| Low vs Very low | -1.330 | 0.176 | 0.264 | 0.187–0.373 | <.001 |
| Medium vs Very low | -2.135 | 0.286 | 0.118 | 0.067–0.207 | <.001 |
| **Area of residence** |  |  |  |  |  |
| HSES vs LSES | -2.005 | 0.177 | 0.135 | 0.095–0.190 | <.001 |

**Note:** $\beta$ = regression coefficient; SE = standard error; OR = odds ratio; CI = confidence interval.

### Health outcomes associated with air pollution exposure

The analysis reveals significant differences in most health outcomes across areas of residence. Respiratory problems were significantly more prevalent in LSES areas (72.7%) compared to HSES areas (50.4%) (*p* < 0.001), with a moderate effect size (Cramér’s V = 0.224). In contrast, allergies, eye irritation, and headaches were all significantly more prevalent in HSES areas, with strong effect sizes (Cramér’s V ranging from 0.429 to 0.508). Specifically, headaches showed the largest disparity (87.0% in HSES vs 38.4% in LSES). No statistically significant difference was observed for cardiovascular conditions (*p* = 0.321), likely due to the very low number of reported cases. The composite health score further indicates that LSES respondents experienced a higher cumulative burden of symptoms, despite lower reporting of some individual conditions. Study results are as shown in Table 7.

**Table 7:** Summary of Health Outcomes by Area of Residence (LSES vs HSES)

| Health Outcome | LSES (n, % Yes) | HSES (n, % Yes) | $\chi^2$ | df | p-value | Cramér's V |
| --- | --- | --- | --- | --- | --- | --- |
| Respiratory Problems | 268 (72.7%) | 194 (50.4%) | 29.081 | 1 | <0.001 | 0.224 |
| Cardiovascular Issues | $\approx 3$ (0.9%) | $n \approx 3$ (0.9%) | 0.985 | 1 | 0.321 | 0.041 |
| Allergies | 68 (18.6%) | 237 (61.7%) | 106.479 | 1 | <0.001 | 0.429 |
| Eye Irritation | 118 (32.2%) | 309 (80.4%) | 136.577 | 1 | <0.001 | 0.486 |
| Headaches | 141 (38.4%) | 334 (87.0%) | 149.180 | 1 | <0.001 | 0.508 |

## DISCUSSION

### Influence of Socioeconomic Factors on Air Quality Exposure

Overall, the results confirm that the two study areas differ significantly across key socioeconomic indicators, including age structure, education, employment, and income. These differences are important in understanding variations in environmental exposure and health outcomes observed in subsequent analyses.

Recent studies have consistently highlighted socioeconomic disparities in urban populations and their implications for environmental exposure and health. A study by Khreis et al. (2020) found that lower socioeconomic groups tend to have lower education levels, higher unemployment rates, and greater exposure to environmental risks. Similarly, research by Brauer et al. (2021) demonstrated that socioeconomic inequalities in urban settings are strongly associated with differences in income, employment, and access to resources, which in turn influence exposure to air pollution.

In the African context, studies conducted in Nairobi informal settlements have shown that residents typically exhibit lower educational attainment, higher unemployment, and lower income levels compared to residents in planned urban areas. These socioeconomic conditions contribute to increased vulnerability to environmental hazards and poorer health outcomes (Muindi et al., 2021). Furthermore, the World Health Organization (2021) emphasizes that socioeconomic status is a key determinant of both environmental exposure and health, with disadvantaged populations facing disproportionate risks due to limited access to resources and services.

The ordinal logistic regression results demonstrate that socioeconomic and residential characteristics were significantly associated with perceived air-quality ratings. Compared with respondents in LSES areas, those residing in HSES areas had significantly lower odds of reporting poorer perceived air quality (OR = 0.135, 95% CI: 0.095–0.190, p < 0.001). Income was also significantly associated with perceived air quality, with respondents earning USD 1–4 per day and USD 5–9 per day having 73.6% and 88.2% lower odds, respectively, of reporting poorer air quality compared with those earning less than USD 1 per day. However, the highest-income category was not statistically significant, likely partly because it contained only five respondents.

Education showed a more limited association. Respondents with no formal education had significantly higher odds of reporting poorer perceived air quality than those with secondary education (OR = 3.254, 95% CI: 1.388–7.638, p = 0.007), while the primary and college/university categories were not statistically significant. Overall, the findings indicate that residential location and income were the strongest predictors of perceived air-quality disparities, supporting the broader evidence that environmental exposures are socially patterned, with socioeconomically disadvantaged communities often experiencing poorer environmental conditions and greater environmental-health risks.

These findings are consistent with previous studies from Nairobi and other settings. In Nairobi, West et al. (2020) documented substantial particulate-matter exposure in an informal settlement and showed that personal exposure varied according to residents’ activities and local environmental conditions. A study by Brauer et al. (2021) found that in many urban African settings, local sources such as cooking emissions, waste burning, and dust significantly contribute to overall PM₂.₅ exposure, often exceeding contributions from industrial activities.

Muindi et al. (2021) reported that residents of informal settlements commonly attribute air pollution to waste burning, road dust, and household fuel use, consistent with the findings of this study. These sources are closely linked to socioeconomic conditions, particularly limited access to clean energy and proper waste disposal systems.

### Health outcomes associated with air pollution exposure

The results demonstrate a complex and nuanced relationship between area of residence and perceived health outcomes. Respiratory problems were significantly more prevalent among respondents living in LSES areas compared to those in HSES areas. This pattern is consistent with the higher exposure to air pollution sources typically found in LSES settings, including open waste burning, use of biomass fuels for cooking, and dust from unpaved roads. These environmental conditions contribute to elevated levels of particulate matter and other pollutants, which are well known to adversely affect respiratory health.

In contrast, allergies, eye irritation, and headaches were reported at substantially higher rates in HSES areas. While this may appear counterintuitive given the generally better environmental conditions in such areas, it likely reflects differences in health awareness, perception, and reporting behavior. Individuals in HSES areas have greater access to healthcare services, higher levels of education, and increased awareness of environmental health risks, leading to more frequent recognition and reporting of symptoms (Binder et al., 2022). Additionally, exposure patterns may differ, with HSES populations potentially experiencing indoor allergens, urban lifestyle-related stressors, or other triggers that contribute to these conditions.

The absence of a statistically significant association between area of residence and cardiovascular conditions is likely attributable to the very low prevalence of reported cases within the sample. Cardiovascular diseases are typically chronic and require clinical diagnosis, making them less likely to be accurately captured through self-reported survey data. This limitation reduces the statistical power to detect meaningful differences between groups.

Recent studies continue to support the observed disparities in health outcomes associated with air pollution exposure. A study by Chang et al. (2023) found that populations living in lower socioeconomic areas experience higher exposure to air pollutants and consequently greater respiratory health risks. Similarly, research by Mohan et al. (2023) demonstrated that particulate matter exposure is strongly associated with increased respiratory morbidity, particularly in urban environments with poor infrastructure.

In the African context, studies conducted in Nairobi and other sub-Saharan cities have shown that residents of informal settlements report higher levels of respiratory symptoms due to exposure to household air pollution and ambient sources such as waste burning and road dust. For instance, Muindi et al. (2021) reported that exposure to particulate matter in Nairobi’s low-income areas is significantly associated with coughing, wheezing, and breathing difficulties. On the other hand, higher reporting of symptoms such as allergies and eye irritation among higher socioeconomic groups has been linked to differences in awareness and healthcare access. A study by Ezzati et al. (2020) noted that individuals with higher socioeconomic status are more likely to recognize and report health symptoms due to better health literacy and access to diagnostic services.

Furthermore, recent evidence indicates that the health impacts of air pollution extend beyond respiratory conditions. According to the World Health Organization (2021), exposure to ambient air pollution contributes to a wide range of health outcomes, including headaches, eye irritation, and long-term cardiovascular effects. However, the latter are often underreported in survey-based studies due to their chronic nature and the need for clinical diagnosis. Overall, these studies reinforce the findings of this research by demonstrating that air pollution exposure and its associated health outcomes are strongly influenced by socioeconomic factors, while also highlighting the role of perception and reporting differences in shaping observed patterns.

## CONCLUSION

This study highlights substantial socioeconomic disparities in perceived air quality and associated health outcomes among residents of Nairobi. Residents in low-socioeconomic status (LSES) areas (Mukuru kwa Njenga) face significantly higher exposure to PM₂.₅ from localized sources such as open waste burning, cooking fuels, and road dust. These areas are characterized by lower educational attainment, higher unemployment, and very low income levels compared to high-socioeconomic status (HSES) areas (Lang’ata). Logistic regression analysis confirmed that area of residence and income are the strongest predictors of perceived air quality, with HSES residence markedly reducing the likelihood of reporting poor air quality. While respiratory problems were more prevalent in LSES areas, certain symptoms like allergies, eye irritation, and headaches were reported more frequently in HSES areas, likely due to higher health awareness.

The study recommends targeted policy interventions to address these environmental injustices. Key recommendations include improving waste management, promoting cleaner cooking fuels, upgrading infrastructure in informal settlements, and enforcing existing air quality regulations more effectively. By tackling these disparities, stakeholders can reduce health inequities and support sustainable urban development in Nairobi and similar cities across Africa.

## Data Availability

The dataset underlying the findings of this study is publicly available in the Zenodo repository at https://doi.org/10.5281/zenodo.21849372.

## ACKNOWGEMENT

We sincerely thank the residents of Langata an Mukuru areas who participated in this study. We are grateful to the community leaders and the research assistants for their support during data collection.

We also wish to thank Willah Nabukwanga, Betty Koech and Joan Kinya of the Kenya Medical Research Institute (Center for Respiratory Diseases Research) for their valuable technical guide and support through the study.

We acknowledge the School of Public Health, Jomo Kenyatta University of Agriculture and Technology, for providing academic and institutional support during this research.

## REFERENCES

Bai, Y., Zhao, T., Hu, W., Zhou, Y., Xiong, J., Wang, Y., … & Zheng, H. (2022). Meteorological mechanism of regional PM2. 5 transport building a receptor region for heavy air pollution over Central China. Science of The Total Environment, 808, 151951.

Begam, G. R., Vachaspati, C. V., Ahammed, Y. N., Kumar, K. R., Babu, S. S., & Reddy, R. R. (2016). Measurement and analysis of black carbon aerosols over a tropical semi-arid station in Kadapa, India. Atmospheric Research, 171, 77–91.

Beguy, D., Elung’ata, P., Mberu, B., Oduor, C., Wamukoya, M., Nganyi, B., & Ezeh, A. (2015). Health & demographic surveillance system profile: the Nairobi urban health and demographic surveillance system (NUHDSS). International journal of epidemiology, 44(2), 462–471.

Brauer, M., Casadei, B., Harrington, R. A., Kovacs, R., Sliwa, K., & WHF Air Pollution Expert Group. (2021). Taking a stand against air pollution—the impact on cardiovascular disease: a joint opinion from the World Heart Federation, American College of Cardiology, American Heart Association, and the European Society of Cardiology. Journal of the American College of Cardiology, 77(13), 1684–1688.

Bravo, M. A., Warren, J. L., Leong, M. C., Deziel, N. C., Kimbro, R. T., Bell, M. L., & Miranda, M. L. (2022). Where is air quality improving, and who benefits? A study of PM2. 5 and ozone over 15 years. American Journal of Epidemiology, 191(7), 1258–1269.

Breysse, P. N., Delfino, R. J., Dominici, F., Elder, A. C., Frampton, M. W., Froines, J. R., … & Wexler, A. S. (2013). US EPA particulate matter research centers: summary of research results for 2005–2011. *Air Quality*, Atmosphere & Health, 6, 333–355.

Brody, S. D., Peck, B. M., & Highfield, W. E. (2004). Examining localized patterns of air quality perception in Texas: A spatial and statistical analysis. Risk Analysis: An International Journal, 24(6), 1561–1574.

Bullard, R. L., Singh, A., Anderson, S. M., Lehmann, C. M., & Stanier, C. O. (2017). 10-Month characterization of the aerosol number size distribution and related air quality and meteorology at the Bondville, IL Midwestern background site. Atmospheric Environment, 154, 348–361.

Chang, J. H., Lee, Y. L., Chang, L. T., Chang, T. Y., Hsiao, T. C., Chung, K. F., … & Chuang, H. C. (2023). Climate change, air quality, and respiratory health: a focus on particle deposition in the lungs. Annals of Medicine, 55(2), 2264881.

Chin, 2019. Public awareness and support for environmental protection – a focus on air pollution in peninsular Malaysia. PLOS One, 14 (3) (2019), Article e0212206.

Clark, L. P., Millet, D. B., & Marshall, J. D. (2014). National patterns in environmental injustice and inequality: outdoor NO2 air pollution in the United States. PloS one, 9(4), e94431.

Cohen, A. J., Brauer, M., Burnett, R., Anderson, H. R., Frostad, J., Estep, K., … & Forouzanfar, M. H. (2017). Estimates and 25-year trends of the global burden of disease attributable to ambient air pollution: an analysis of data from the Global Burden of Diseases Study 2015. The lancet, 389(10082), 1907–1918.

Cromar, K. R., Gladson, L. A., Perlmutt, L. D., Ghazipura, M., & Ewart, G. W. (2016). American Thoracic Society and Marron Institute report. Estimated excess morbidity and mortality caused by air pollution above American Thoracic Society–recommended standards, 2011– 2013. Annals of the American Thoracic Society, 13(8), 1195–1201.

Damette, O., Delacote, P., & Del Lo, G. (2018). Households energy consumption and transition toward cleaner energy sources. Energy Policy, 113, 751–764.

Deng, Z., Chen, F., Zhang, M., Lan, L., Qiao, Z., Cui, Y., … & Li, X. (2016). Association between air pollution and sperm quality: a systematic review and meta-analysis. Environmental pollution, 208, 663–669.

deSouza, P., & Kinney, P. L. (2021). On the distribution of low-cost PM2. 5 sensors in the US: demographic and air quality associations. Journal of exposure science & environmental epidemiology, 31(3), 514–524.West et al. (2020)

Du, W., Shen, G., Chen, Y., Zhu, X., Zhuo, S., Zhong, Q., … & Tao, S. (2017). Comparison of air pollutant emissions and household air quality in rural homes using improved wood and coal stoves. Atmospheric Environment, 166, 215–223.

Egondi, T., Muindi, K., Kyobutungi, C., Gatari, M., & Rocklöv, J. (2016). Measuring exposure levels of inhalable airborne particles (PM2. 5) in two socially deprived areas of Nairobi, Kenya. Environmental research, 148, 500–506.Ngo et al. (2018

Geng, G., Xiao, Q., Liu, S., Liu, X., Cheng, J., Zheng, Y., … & Zhang, Q. (2021). Tracking air pollution in China: near real-time PM2. 5 retrievals from multisource data fusion. Environmental Science & Technology, 55(17), 12106–12115.

Gong, K., Li, L., Li, J., Qin, M., Wang, X., Ying, Q., … & Hu, J. (2021). Quantifying the impacts of inter-city transport on air quality in the Yangtze River Delta urban agglomeration, China: Implications for regional cooperative controls of PM2. 5 and O3. Science of the Total Environment, 779, 146619.

Grunig, G., Marsh, L. M., Esmaeil, N., Jackson, K., Gordon, T., Reibman, J., … & Park, S. H. (2014). Perspective: ambient air pollution: inflammatory response and effects on the lung’s vasculature. Pulmonary circulation, 4(1), 25–35.

Gulis, V., Rosemond, A. D., Suberkropp, K., Weyers, H. S., & Benstead, J. P. (2004). Effects of nutrient enrichment on the decomposition of wood and associated microbial activity in streams. Freshwater Biology, 49(11), 1437–1447.

Guttikunda, S. K., Nishadh, K. A., & Jawahar, P. (2019). Air pollution knowledge assessments (APnA) for 20 Indian cities. Urban Climate, 27, 124–141.

Han, C., Lim, Y. H., Yorifuji, T., & Hong, Y. C. (2018). Air quality management policy and reduced mortality rates in Seoul Metropolitan Area: A quasi-experimental study. Environment international, 121, 600–609.

Health Effects Institute (2020). State of Global Air 2020. Special Report. Boston, MA:Health Effects Institute.

Hicken, M. T., Payne-Sturges, D., & McCoy, E. (2023). Evaluating Race in Air Pollution and Health Research: Race, PM2. 5 Air Pollution Exposure, and Mortality as a Case Study. Current Environmental Health Reports, 1–11.

Ho, H. C., Wong, P. P., & Guo, C. (2021). Impacts of social and environmental perceptions on preparedness and knowledge of air pollution risk: A study of adolescent males in an urbanized, high-density city. Sustainable Cities and Society, 66, 102678.

Howse, E., Crane, M., Hanigan, I., Gunn, L., Crosland, P., Ding, D., … & Rychetnik, L. (2021). Air pollution and the noncommunicable disease prevention agenda: opportunities for public health and environmental science. Environmental Research Letters, 16(6), 065002.

Huang, Z., Yu, Q., Ma, W., & Chen, L. (2019). Surveillance efficiency evaluation of air quality monitoring networks for air pollution episodes in industrial parks: Pollution detection and source identification. Atmospheric Environment, 215, 116874.

Jing, H. (2015, January). Empirical Study on the Urbanization Promoting the Chinese Economic Growth. In 2014 International Conference on Computer Science and Electronic Technology (ICCSET 2014) (pp. 417–420). Atlantis Press.

Johnson, F., Kanninen, B., Bingham, M., & Özdemir, S. (2007). Experimental design for stated-choice studies. Valuing environmental amenities using stated choice studies, 159–202.

Kenya National Bureau of Statistics (2022). Economic Survey.

Khreis, H., Nieuwenhuijsen, M. J., Zietsman, J., & Ramani, T. (2020). Traffic-related air pollution: Emissions, human exposures, and health: An introduction. In Traffic-related air pollution (pp. 1–21). Elsevier.

Lamichhane, D. K., Lee, S. Y., Ahn, K., Kim, K. W., Shin, Y. H., Suh, D. I., … & Kim, H. C. (2020). Quantile regression analysis of the socioeconomic inequalities in air pollution and birth weight. Environment International, 142, 105875.

Landrigan, P. J. (2017). Air pollution and health. The Lancet Public Health, 2(1), e4–e5.

Landrigan, P. J. (2017). Air pollution and health. The Lancet Public Health, 2(1), e4–e5.

Liao, P. S., Shaw, D., & Lin, Y. M. (2015). Environmental quality and life satisfaction: Subjective versus objective measures of air quality. Social Indicators Research, 124, 599–616.

Lin, B., & Zhu, J. (2018). Changes in urban air quality during urbanization in China. Journal of Cleaner Production, 188, 312–321.

Liu, 2016. Public’s health risk awareness on urban air pollution in Chinese megacities: The cases of Shanghai, Wuhan and Nanchang. Int. J. Environ. Res. Public Health, 13 (9) (2016), p. 845.

Maantay, J. (2007). Asthma and air pollution in the Bronx: methodological and data considerations in using GIS for environmental justice and health research. Health & place, 13(1), 32–56.

Mberu, B., Wamukoya, M., Oti, S., & Kyobutungi, C. (2015). Trends in causes of adult deaths among the urban poor: evidence from Nairobi urban health and demographic surveillance system, 2003–2012. Journal of Urban Health, 92, 422–445.

Mohan, A., Alupo, P., Martinez, F. J., Mendes, R. G., Zhang, J., & Hurst, J. R. (2023). Respiratory health and cities. American journal of respiratory and critical care medicine, 208(4), 371–373.

Mueller, S., Tarnay, L., O’Neill, S., & Raffuse, S. (2020). Apportioning smoke impacts of 2018 wildfires on eastern Sierra Nevada sites. Atmosphere, 11(9), 970.

Murray, N. L., Holmes, H. A., Liu, Y., & Chang, H. H. (2019). A Bayesian ensemble approach to combine PM2. 5 estimates from statistical models using satellite imagery and numerical model simulation. Environmental research, 178, 108601.

Naidja, L., Ali-Khodja, H., & Khardi, S. (2017). Particulate matter from road traffic in Africa. Journal of Earth Sciences and Geotechnical Engineering, 7(1), 389–304.

Nikolopoulou, M., Kleissl, J., Linden, P. F., & Lykoudis, S. (2011). Pedestrians’ perception of environmental stimuli through field surveys: Focus on particulate pollution. Science of the total environment, 409(13), 2493–2502.

Odonkor, S. T., & Mahami, T. (2020). Knowledge, attitudes, and perceptions of air pollution in Accra, Ghana: a critical survey. Journal of environmental and public health, 2020.

Oudin, A., Forsberg, B., Adolfsson, A. N., Lind, N., Modig, L., Nordin, M., … & Nilsson, L. G. (2016). Traffic-related air pollution and dementia incidence in northern Sweden: a longitudinal study. Environmental health perspectives, 124(3), 306–312.

Pantavou, K., Lykoudis, S., & Psiloglou, B. (2017). Air quality perception of pedestrians in an urban outdoor Mediterranean environment: A field survey approach. Science of the total environment, 574, 663–670.

Park, S., Mossmann, D., Chen, Q., Wang, X., Dazert, E., Colombi, M., … & Hall, M. N. (2022). Transcription factors TEAD2 and E2A globally repress acetyl-CoA synthesis to promote tumorigenesis. Molecular cell, 82(22), 4246–4261.

Peled, R. (2011). Air pollution exposure: Who is at high risk?. Atmospheric Environment, 45(10), 1781–1785.

Peng, W., Yang, J., Lu, X., & Mauzerall, D. L. (2018). Potential co-benefits of electrification for air quality, health, and CO2 mitigation in 2030 China. Applied energy, 218, 511–519.Mohammed et al. 2023

Penza, M., Suriano, D., Villani, M. G., Spinelle, L., & Gerboles, M. (2014, November). Towards air quality indices in smart cities by calibrated low-cost sensors applied to networks. In SENSORS, 2014 IEEE (pp. 2012–2017). IEEE.Sun et al., 2016

Pu, S., Shao, Z., Fang, M., Yang, L., Liu, R., Bi, J., & Ma, Z. (2019). Spatial distribution of the public’s risk perception for air pollution: A nationwide study in China. Science of the Total Environment, 655, 454–462.

Rotko, T., Oglesby, L., Künzli, N., Carrer, P., Nieuwenhuijsen, M. J., & Jantunen, M. (2002). Determinants of perceived air pollution annoyance and association between annoyance scores and air pollution (PM2. 5, NO2) concentrations in the European EXPOLIS study. Atmospheric Environment, 36(29), 4593–4602.

Schraufnagel, D. E., Balmes, J. R., Cowl, C. T., De Matteis, S., Jung, S. H., Mortimer, K., … & Wuebbles, D. J. (2019). Air pollution and noncommunicable diseases: A review by the Forum of International Respiratory Societies’ Environmental Committee, Part 2: Air pollution and organ systems. Chest, 155(2), 417–426.

Sun, C., Luo, Y., & Li, J. (2018). Urban traffic infrastructure investment and air pollution: Evidence from the 83 cities in China. Journal of cleaner production, 172, 488–496.

World Health Organization. (2014). WHO guidelines for indoor air quality: household fuel combustion. World Health Organization.

World Health Organization. (2021). WHO global air quality guidelines: particulate matter (PM2. 5 and PM10), ozone, nitrogen dioxide, sulfur dioxide and carbon monoxide. World Health Organization.

Yang, X., Ruby Leung, L., Zhao, N., Zhao, C., Qian, Y., Hu, K., … & Chen, B. (2017). Contribution of urbanization to the increase of extreme heat events in an urban agglomeration in east China. Geophysical Research Letters, 44(13), 6940–6950.

Zeng, D. Z., & Zhao, L. (2009). Pollution havens and industrial agglomeration. Journal of Environmental Economics and Management, 58(2), 141–153.

Zhang, Q., Zheng, Y., Tong, D., Shao, M., Wang, S., Zhang, Y., … & Hao, J. (2019). Drivers of improved PM2. 5 air quality in China from 2013 to 2017. Proceedings of the National Academy of Sciences, 116(49), 24463–24469.S

